# SARS-CoV-2 seropositivity and subsequent infection risk in healthy young adults: a prospective cohort study

**DOI:** 10.1101/2021.01.26.21250535

**Authors:** Andrew G. Letizia, Yongchao Ge, Sindhu Vangeti, Carl Goforth, Dawn L Weir, Natalia A. Kuzmina, Hua Wei Chen, Dan Ewing, Alessandra Soares-Schanoski, Mary-Catherine George, William D. Graham, Franca Jones, Preeti Bharaj, Rhonda A. Lizewski, Stephen A. Lizewski, Jan Marayag, Nada Marjanovic, Clare Miller, Sagie Mofsowitz, Venugopalan D. Nair, Edgar Nunez, Danielle M. Parent, Chad K. Porter, Ernesto Santa Ana, Megan Schilling, Daniel Stadlbauer, Victor Sugiharto, Michael Termini, Peifang Sun, Russell. P. Tracy, Florian Krammer, Alexander Bukreyev, Irene Ramos, Stuart C. Sealfon

**Affiliations:** Naval Medical Research Center, Silver Spring, MD USA; Department of Neurology. Icahn School of Medicine at Mount Sinai, New York, NY USA; Department of Pathology University of Texas Medical Branch and Galveston National Laboratory, Galveston, TX USA; Naval Medical Research Unit SIX, Lima Peru; Department of Pathology & Laboratory Medicine, Larner College of Medicine, University of Vermont, Burlington, VT USA; Department of Microbiology, Icahn School of Medicine at Mount Sinai, New York, NY USA; Naval Medical Readiness and Training Command Beaufort, Beaufort, SC USA

**Author notes:** Correspondence to: Dr. Stuart C. Sealfon Icahn School of Medicine at Mount Sinai, Annenberg 14-44, One Gustave L. Levy Place, New York, NY 10029 USA. Contributed equally (Andrew G. Letizia, Yongchao Ge, Sindhu Vangeti, Carl Goforth, Dawn L Weir).

## Abstract

**Background:** The risk of severe acute respiratory syndrome coronavirus 2 (SARS-CoV-2) subsequent infection among seropositive young adults was studied prospectively.

**Methods:** The study population comprised 3,249 predominantly male, 18-20-year-old Marine recruits. Upon arrival at a Marine-supervised two-week quarantine, participants were assessed for baseline SARS-CoV-2 IgG seropositivity, defined as a 1:150 dilution or greater on receptor binding domain and full-length spike protein enzyme-linked immunosorbent (ELISA) assays. SARS-CoV-2 infection was assessed by PCR at initiation, middle and end of the quarantine. After appropriate exclusions, including participants with a positive PCR during quarantine, we performed three biweekly PCR tests in both seropositive and in seronegative groups once recruits left quarantine and entered basic training and baseline neutralizing antibody titers on all subsequently infected seropositive and selected seropositive uninfected participants.

**Findings:** Among 189 seropositive participants, 19 (10.1%) had at least one positive PCR test for SARS-CoV-2 during the six-week follow-up (1.1 cases per person-year). In contrast, 1,079 (48.0%) of the 2,247 seronegative participants tested positive (6.2 cases per person-year). The incidence rate ratio was 0.18 (95% CI 0.11-0.28, p<0.00001). Among seropositive recruits, infection was associated with lower baseline full-length spike protein IgG titers (p<0.0001). Compared with seronegative recruits, seropositive recruits had about 10-fold lower viral loads (ORF1ab gene, p<0.005), and trended towards shorter duration of PCR positivity (p=0.18) and more frequent asymptomatic infections (p=0.13). Among seropositive participants, baseline neutralizing titers were detected in 45 of 54 (83.3%) uninfected and in 6 of 19 (31.6%) infected participants during the 6 weeks of observation (ID50 difference p<.0001).

**Interpretation:** Seropositive young adults had about one-fifth the risk of subsequent infection compared with seronegative individuals. Although antibodies induced by initial infection are largely protective, they do not guarantee effective SARS-CoV-2 neutralization activity or immunity against subsequent infection. These findings may be relevant for optimization of mass vaccination strategies.

**Funding:** Defense Health Agency and Defense Advanced Research Projects Agency

## Introduction

As of mid-December 2020, more than 72 million severe acute respiratory syndrome coronavirus 2 (SARS-CoV-2) infections have been diagnosed world-wide^1^. Serological surveys indicate that the actual number of infections has been many times higher than the cumulative incidence of diagnosed cases, with seropositivity rates approaching 10% in some countries and more than 40% in the Brazilian Amazon^2-4^. With the onset of mass SARS-CoV-2 vaccination programs and the increasing proportion of previously infected individuals, the risk of reinfection after natural infection is an important question for modeling the pandemic, estimating herd immunity and guiding vaccination strategies^5,6^.

Most individuals mount a sustained serologic response after initial infection^7-11^. Similar to the response to other coronaviruses, the production of IgG antibodies against SARS-CoV-2 peaks several weeks after infection, goes through a decline phase and then stabilizes. The overall humoral response to SARS-CoV-2 is highly variable among individuals^9^. SARS-CoV-2 specific IgG antibodies can be detected in serum from most individuals several months after infection^11^. However, a percentage of infected patients, ranging from 2.5% to 28% in different studies, do not maintain detectable circulating antibodies^9^ or neutralizing activity^10-12^ at later time points. About 10% of individuals who developed antibodies to SARS-CoV-2, all of whom had full-length spike protein-specific IgG antibody titers lower than 1:320, failed to develop measurable neutralizing activity^11^.

Reports have established that SARS-CoV-2 reinfection occurs after previous infection as well as in seropositive individuals^13-22^. Several studies have reported that SARS-CoV-2 IgG antibodies^22,23^ and neutralizing antibodies^24^ provide protection against subsequent infection. The initial trial results of the adenoviral vector-based SARS-CoV-2 vaccine, ChAdOx1 nCoV-19 (AZD1222), noted that among the 373 participants who were seropositive at baseline, 3 (0.8%) had subsequent positive swab PCR tests; in comparison, among 11,263 baseline seronegative participants in all arms of that study, 218 (1.9%) developed a positive test^21^. A study of SARS-CoV-2 serological status and infection among healthcare workers identified 2 infections among 1265 (0.16%) who were seropositive and 223 among 11,364 (2.0%) who were seronegative^22^. Young adults, of whom a high proportion are asymptomatically infected and become seropositive in the absence of known infection^25,26^ can be an important source of transmission to more vulnerable populations^27^. Evaluating the protection against subsequent SARS-CoV-2 infection conferred by seropositivity in young adults is important for determining the need for vaccinating previously infected individuals in this age group.

We utilized the COVID-19 Health Action Response for Marines (CHARM) study^26^, a longitudinal prospective cohort study, to examine the effect of SARS-CoV-2 seropositivity on the risk of developing SARS-CoV-2 infection in young, healthy adult Marine recruits.

## Methods

### Study design and participants

The prospective study observation period began when Marine recruits arrived at Marine Corps Recruit Depot – Parris Island (MCRDPI) to commence basic training (Figure 1). To mitigate the spread of SARS-CoV-2, just before transferring to MCRDPI, the United States Marine Corps (USMC) implemented two separate quarantine protocols. The first was a two-week home quarantine. After that, the recruits traveled, while masked and socially distanced, to a second USMC-supervised quarantine situated at either a college campus from May until July 2020 or at a hotel from August until October 2020. The supervised quarantine employed extensive public health measures, as previously described^26^, that were strictly enforced by US Marine instructors at all times. The recruits and staff were forbidden to leave, and no visitors other than deliveries of supplies and food along with local essential workers and the study staff were allowed onto the premises. At the end of this quarantine period, the USMC required all recruits to test negative for SARS-CoV-2 by PCR before proceeding to MCRDPI to initiate basic training.

**Figure 1:**
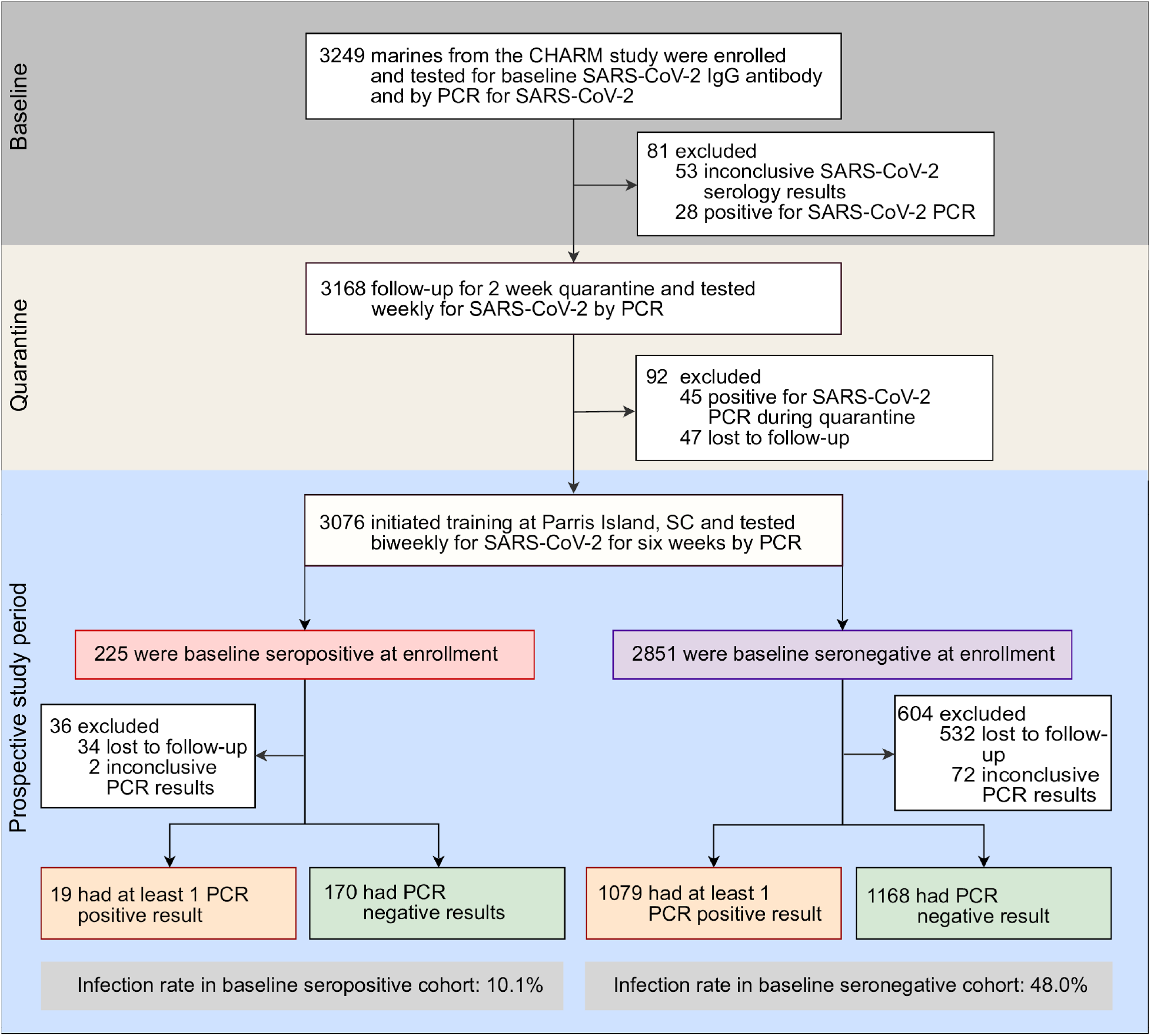
Flow chart of the study design and outcomes. The baseline analyses were performed within two days of enrollment and two weeks before the prospective study period. Nearly all participants were tested at scheduled biweekly intervals during the prospective study period. A few participants were diagnosed by the Marine Corps Recruit Depot Parris Island clinic, and were tested by the study team at times not corresponding to the regularly scheduled longitudinal follow ups. Participants lost to follow up either dropped out of the study, were separated from the Marine Corps or were removed from the base for medical or administrative reasons. The study team did not know the reason for participants missing study visits.

Within 48 hours of arriving at the supervised quarantine location, recruits were offered the opportunity to volunteer for CHARM. Recruits were eligible if they were ≥18 years of age. Since recruits are a vulnerable population and at risk for coercion, special measures were undertaken including study briefers, who are active-duty Navy personnel wore civilian clothes, did not disclose military ranks, did not have members in the recruit’s chain of command present, and ensured that participation would not affect a recruit’s medical care or influence the grading of a recruit’s military performance by superiors. Institutional Review Board approval was obtained from the Naval Medical Research Center (protocol number NMRC.2020.0006) in compliance with all applicable U.S. federal regulations governing the protection of human subjects. All participants provided written informed consent.

At enrollment, participants completed a questionnaire consisting of demographic information, risk-factors, reporting of 14 specific COVID-19 related symptoms or any other unspecified symptom, and brief medical-history. At quarantine weeks 0, 1 and 2 a mid-turbinate nares swab for SARS-CoV-2 PCR testing and sera were obtained and questionnaire administered. The follow up questionnaire inquired about the same COVID-19 related symptoms since the last study visit.

Participants who had three negative swab PCR results at the beginning, middle and end of quarantine and a baseline serum serology test at the beginning of the supervised quarantine that identified them as seronegative or seropositive for SARS-CoV-2 according to criteria described below were followed prospectively. At weeks 2, 4 and 6 after transfer from the quarantine location to MCRDPI, a mid-turbinate nares swab for SARS-CoV-2 PCR testing and sera were obtained and the symptom questionnaire administered. When clinically indicated due to the development of symptoms, some participants were evaluated at the MCRDPI clinic and diagnosed by rapid testing. If positive, they went to the isolation barracks, where the study team was able to follow up and repeat testing outside of the scheduled longitudinal follow up encounters. Participants without PCR results obtained during the MCRDPI study period were excluded from analysis.

### Procedures

#### SARS-CoV-2 quantitative PCR testing

All swabs in viral transport media (VTM) were kept at 4°C. All assays were performed within 48h of sample collection at high complexity Clinical Laboratory Improvement Amendments (CLIA)-certified laboratories using the US FDA authorized Thermofisher TaqPath™ COVID-19 Combo Kit (Thermo Fisher Scientific, Waltham MA). Lab24Inc (Boca Raton, FL) performed PCR testing from study initiation until the end of July, and the Naval Medical Research Center (Silver Spring, MD) from August until the conclusion of the study.

#### SARS-CoV2 enzyme-linked immunosorbent assay (ELISA)

The presence and levels of IgG SARS-CoV-2-specific antibodies in serum were determined using an enzyme-linked immunosorbent assay (ELISA) as previously described^26^. Briefly, 384-well Immulon 4 HBX plates (Thermofisher), or 96-well half area Microlon plates (Greiner Bio-One), were coated overnight at 4°C with recombinant his-tagged spike (S) receptor binding domain (S-RBD) (SinoBiological) or full-length spike protein (LakePharma) at a concentration of 2 µg/ml in phosphate-buffered saline (PBS). Plates were washed three times with 0.1% Tween-20 (Fisher Scientific) in PBS (PBS-T) using an automated ELISA plate washer (Aquamax 4000, Molecular devices), and blocked for 1 h at room temperature (RT) with 3% milk (BioRad) PBS-T. Blocking solution was removed, and serum samples diluted in 1% milk PBS-T were dispensed in the wells. At least 2 positive controls (sera with known IgG presence), 8 negative controls (sera collected before July 2019) and 4 blanks (no serum) were included in every assay. Plates were incubated for 2 h at room temperature, and then washed 3 times with PBS-T. Next, peroxidase conjugated goat F(ab’)2 Anti-Human IgG (Abcam) was added at a dilution 1:5,000-1:10,000 dilutions (determined after optimization for each antibody lot) in 1% milk PBS-T, and plates were incubated for 1 h at room temperature. Plates were washed 6 times with PBS-T, developed using o-Phenylenediamine (Sigma), and the reaction was stopped after 10 min with 3M HCl. Optical density (OD) at 492 nm was measured using a microplate reader (SpectramaxM2, Molecular Devices). All serum samples were screened at a 1:50 dilution with S-RBD. Those samples with an OD 492 nm value higher than the average of the negative controls plus 3 times their standard deviation (SD) in the screening assay underwent titration assay (6 serial 1:3 serum dilutions starting at 1:50) using both S-RBD and full-length spike protein. Serum samples were considered positive for each assay when at least 2 consecutive dilutions showed higher OD 492 nm than the average of the negative controls plus 3 times their SD at the correspondent dilution or 0.15 OD 492 nM. Specificity was 100% on both S-RBD and full-length spike protein ELISA using 70 control sera obtained before July 2019. Participants were only considered seropositive to SARS-CoV-2 if IgG titrations for both ELISA gave a positive result at a minimum of 1:150 dilution.

#### Neutralization assays

Two-fold serial dilutions of heat-inactivated serum at an initial dilution of 1:20, were prepared in serum free media (Minimum Essential Medium; Thermo fisher Scientific, Cat No. 11095080 containing 25 mM HEPES and 0.05 g/L gentamicin sulfate) and incubated with an equal volume of mNeonGreen SARS-CoV-2^28^ for 1 hour at 37°C at a final concentration of 200 plaque forming units in humidified 5% CO_2_. Virus-serum mixtures were then added to Vero-E6 monolayers in 96 well optical black plates and incubated at 37°C. Plates were read using the BioTek Cytation 5 plate reader (EX 485 nm, EM 528 nm) at 24 h post-infection. Following background signal correction, neutralization titers at a fluorescent end point of 50% virus reduction (ID_50_) were determined.

#### Statistical Analysis

Analyses, figures and tables were generated using R 3.6.3. Race was categorized as non-Hispanic white, non-Hispanic black, non-Hispanic other, and Hispanic. Cochran-Armitage Chi-square test for trend was used to compare proportions testing positive for SARS-CoV-2 by increasing titers. Cumulative incidence rates computed by Kapan-Meir method were used to estimate the risk of SARS-CoV-2 infection between seropositive and seronegative participants and also between different titers among the seropositive participants. Observational follow-up began upon arrival at MCRDPI and participants were censored at the first observed positive PCR, or at the latest time with valid PCR assay or at the termination of the study (6 weeks of observation). The Cox proportional hazards model controlled for age, sex and race. The p-values from the cumulative incidence curves were determined by the log-rank test and the p-value and CI for Cox proportional hazard model was computed by the R function coxph. The cycle threshold (Ct) values of viral genes were quantile normalized to remove the batch effects between Lab24Inc and Naval Medical Research Center laboratory assays. The CI and p-values for comparing the mean of the Ct values for two groups are computed based on the two sample t-test.

## Results

The flow of participants through the study is shown in Figure 1. Two weeks before the initiation of basic training and the start of the prospective cohort study period, a total of 3,249 out of 4,657 eligible new Marine recruits (70%) enrolled in CHARM and underwent a supervised two-week quarantine. Exclusions before the prospective study period included 73 participants who were SARS-CoV-2 PCR positive on at least one of the three PCR tests performed during quarantine, 53 who lacked baseline serology results and 47 who were lost to follow-up.

Of the remaining 3,076 participants, 225 were baseline seropositive, having SARS-CoV-2 IgG titers in serum samples obtained at the beginning of quarantine that were greater than 1:150 both with S-RBD and with full-length spike protein ELISA, and 2,851 were baseline seronegative. Among the seropositive participants, 36 (16.0%) were excluded from analysis due either to being lost to follow up (n=34), or to lacking valid PCR results during the study period (n=2). In the seronegative group, 604 (21.2%) participants were excluded due to either being lost to follow up (n=532), or lacking valid PCR results (n=72). Participants were lost to follow up for specific reasons unknown to the study team, including dropping out of the study, being separated from the Marines or being removed from the base for medical or administrative reasons. Demographic characteristics of the two groups in the study cohort are shown in Table 1. Most participants were 18-20 years old and male. The two groups were well-balanced with the exception of a higher proportion of participants who self-identified as Hispanic and as black in the seropositive group.

**Table 1:**
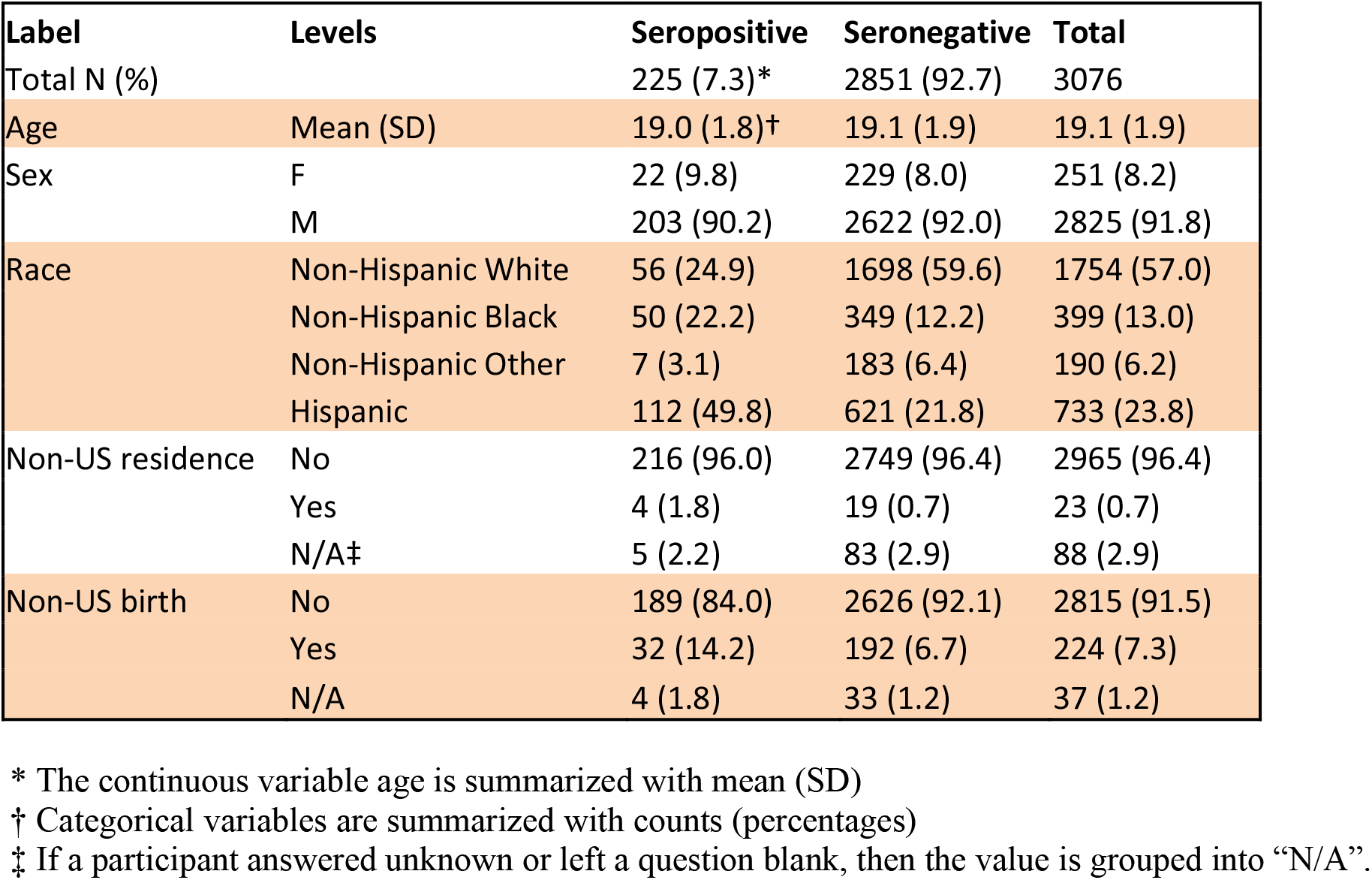
Demographic characteristics of participants studied prospectively in the seropositive and seronegative groups. A total of 120 participants were excluded due to being lost to follow up, lacking any baseline valid IgG or becoming PCR positive during the quarantine period. The table includes all 3,076 participants who entered training and were followed prospectively, including the 640 participants who were later excluded for further analysis (see Figure 1).

A total of 19 out of 189 (10.1%, 1.1 cases per person-year) seropositive participants and 1,079 out of 2,247 (48.0%, 6.2 cases per person-year) seronegative participants had at least one positive SARS-CoV-2 PCR result during the six-week study period, representing an 0.18 (95% CI: 0.11 to 0.28, p<0.001) incidence rate ratio for SARS-CoV-2 infections in the seropositive group (Table 2). The temporal incidence of infection in seropositive and seronegative groups is shown in Figure 2A. After adjusting the effects of race, age and sex on the SARS-CoV-2 infections, the hazard ratio comparing seropositive participants and seronegative participants was 0.16 (95% CI, 0.10-0.25, p<0.001; supplementary Table 1).

**Table 2:** Comparison of SARS-Cov-2 infection (PCR positive) at MCRDPI between the seropositive and seronegative groups. The analysis is based on the 2,436 participants who had valid PCR data obtained during the prospective follow-up period.

| Group | Seropositive | Seronegative |
| --- | --- | --- |
| Total | 189 | 2247 |
| PCR positive | 19 | 1079 |
| Incidence proportion (%) | 10.1 | 48 |
| Proportion difference (%) | -38 (-45 to -31, p<0.001) |  |
| Observed person year | 17.1 | 175.2 |
| Incidence rate | 1.11 | 6.16 |
| Incidence rate ratio | 0.18 (0.11 to 0.28, p<0.001) |  |

**Figure 2:**
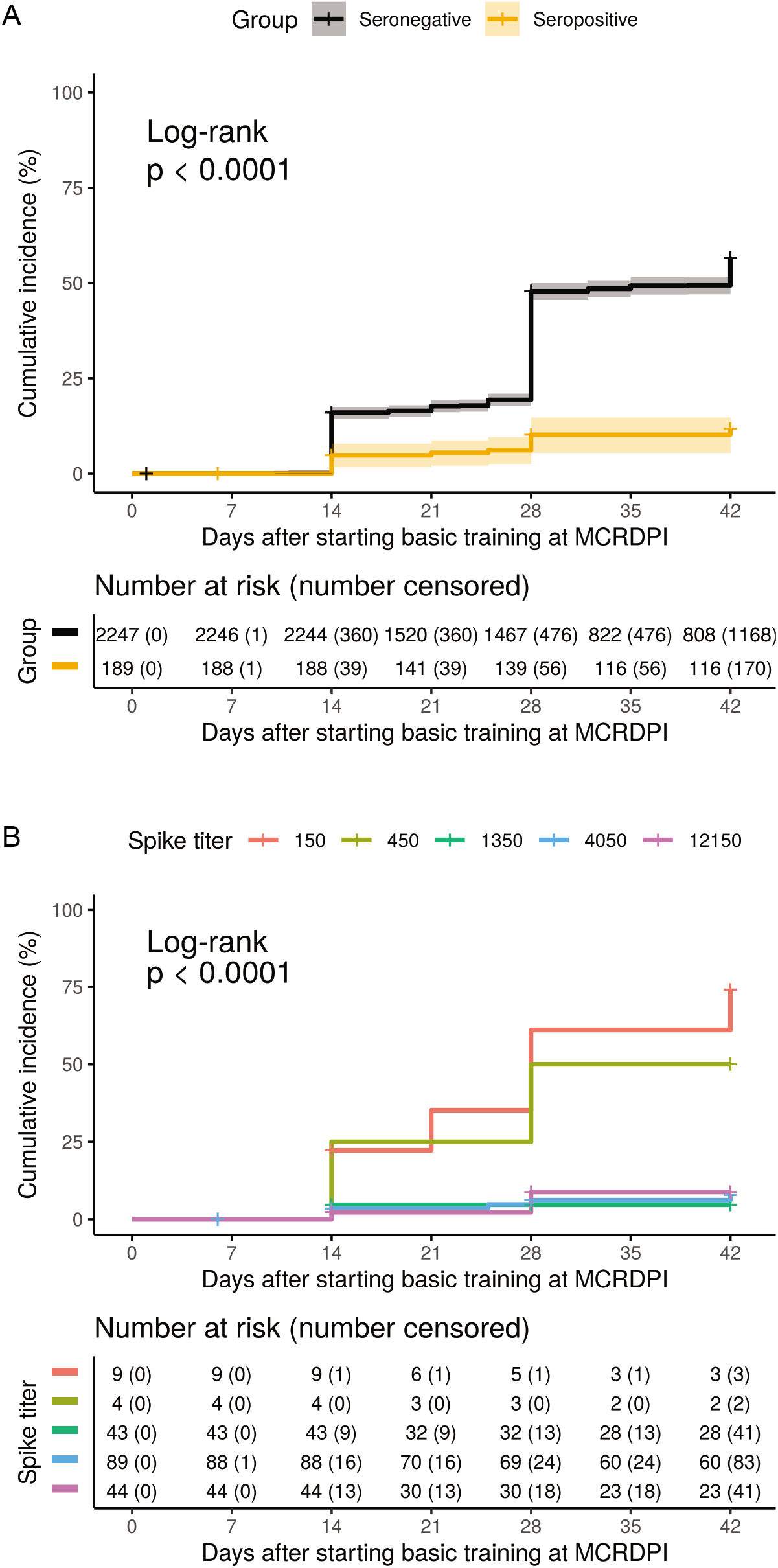
SARS-CoV-2 PCR positive incidence curves during the six-week follow-up period. A. Overall cumulative incidence for testing PCR positive in the baseline seropositive and seronegative groups. B. Cumulative incidence for testing PCR positive in the seropositive group at different baseline full-length spike protein IgG titers which ranged from 1:150 to 1:12150. The cumulative incidence rate is computed by the Kaplan-Meier method, and the cumulated censored participants are listed.

Within the seropositive group, we assessed the association between the SARS-CoV-2 IgG baseline titers and the risk of infection. As shown in Figure 2B, supplementary Figure 2 and Table 3, we found a strong association between subsequent PCR positive infection and lower titers of IgG antibodies directed to full-length spike protein (p<0.0001) as well as to S-RBD (p=0.0019). The detailed Cox proportional hazard analysis in supplementary Table 1 gives a hazard ratio of 0.45 (95% CI 0.32-0.65, p<0.001) and 0.67 (95% CI 0.47-0.96, p=0.028) respectively for the full-length spike protein titer and S-RBD titer, both log-transformed.

**Table 3.** SARS-CoV-2 S-RBD and full-length spike IgG titers and neutralizing antibody activity in PCR positive and PCR negative seropositive participants. p-values were assessed by the Cochran-Armitage test. ID50 is the titer at which a 50% reduction in virus infection was observed.

| Assay | Group | N | No. (%) of people at the titer or ID50 interval |  |  |  |  | mean±sem | p-value |
| --- | --- | --- | --- | --- | --- | --- | --- | --- | --- |
| titer |  |  | 150 | 450 | 1350 | 4050 | 12150 |  |  |
| S-RBD | PCR+ | 19 | 5(26.3) | 5(26.3) | 6(31.6) | 3(15.8) | 0(0) | 674.5±181.1 | 0.017 |
|  | PCR- | 170 | 6(3.5) | 49(28.8) | 81(47.6) | 27(15.9) | 7(4.1) | 1186.3±86.2 |  |
| Full-length spike protein | PCR+ | 19 | 6(31.6) | 2(10.5) | 2(10.5) | 6(31.6) | 3(15.8) | 1202.6±472.7 | 1.20E-05 |
|  | PCR- | 170 | 3(1.8) | 2(1.2) | 41(24.1) | 83(48.8) | 41(24.1) | 3723.7±259.9 |  |
| ID50 range |  |  | <20 | [20,40) | [40,80) | [80,160) | [160,320) |  |  |
| Neutralization | PCR+ | 19 | 13(68.4) | 2(10.5) | 3(15.8) | 1(5.3) | 0(0) | 16.8±3.1* | 9.80E-05 |
|  | PCR- | 54 | 9(16.7) | 10(18.5) | 19(35.2) | 15(27.8) | 1(1.9) | 48.2±5.7 |  |

\*The mean and SEM is computed after converting undetectable titer <20 to be 10.

We examined baseline SARS-CoV-2 IgG neutralizing antibody activity in all seropositive participants who became PCR positive during the observation period and in the first 54 participants who were seropositive but remained PCR negative. Neutralizing activity was above the limit of detection in 45 of 54 (83.3%) seropositive participants who never became PCR positive and in 6 of 19 (31.6%) of participants infected during the 6 weeks of observation. The neutralizing activity assessed as 50% inhibitory dose (ID50) was significantly higher in the participants who did not become PCR positive during the study (p<.0001, Cochran-Armitage test, Table 3, supplementary Figure 2).

We also compared virus loads estimated by PCR Ct values between the seronegative and seropositive PCR infected groups and found that seronegative individuals had on average 4, 2.6 and 3.3 lower cycle values for ORF1ab gene (p=0.004, 95% CI 1.23 to 6.67), S gene (p=0.11, 95% CI -0.58 to 5.77) and N gene (p=0.033, 95% CI 0.27 to 6.33), respectively, than seropositive individuals. The lower Ct values suggest an approximately 10-fold higher virus load in the samples from seronegative participants (Table 4). In addition, seronegative participants tended to have a longer duration of PCR positivity than seropositive individuals (p=0.18). The proportion of asymptomatic participants was 84.2% (16 of 19) and 67.8% (732 of 1079) in seropositive and seronegative infected individuals, respectively, a difference that did not reach significance (p=0.13).

**Table 4:** Comparison of symptoms and cycle threshold values between SARS-CoVCov-2 infected (PCR positive) seropositive and seronegative groups.

|  | Seropositive | Seronegative | Difference comparison |
| --- | --- | --- | --- |
| **Total N** | **19** | **1079** |  |
| Infection duration>7 days | 6 (31.6%)* | 510 (47.3%) | -0.16 (-0.38 to 0.07, p=0.175)† |
| Symptomatic | 3 (15.8%) | 347 (32.2%) | -0.16 (-0.38 to 0.05, p=0.129) |
| N gene Ct | 27.7 (7.6) ‡ | 24.4 (5.5) | 3.30 (0.27 to 6.33, p=0.033) ¶ |
| S gene Ct | 26.9 (7.1) | 24.3 (5.3) | 2.60 (-0.58 to 5.77, p=0.109) |
| ORF1ab Ct | 28.0 (7.0) | 24.0 (5.3) | 3.95 (1.23 to 6.67, p=0.004) |

\* Binary variables are summarized with counts (percentages)
† Difference in proportion for binary variables
‡ The cycle threshold (Ct) values of three viral genes are summarized by mean (SD)
¶ Difference in mean for the Ct values

## Discussion

This study of primarily young, male Marine recruits found that the presence of antibodies to SARS-CoV-2 conferred an 82% reduced incidence rate of SARS-CoV-2 infection. The percentage of symptomatic infections was comparable in seropositive and seronegative participants. Among the seropositive group, the participants that became infected had lower antibody titers than those that were uninfected, and were more likely to lack detectable baseline neutralizing antibody activity. Our results indicate that although antibodies induced by infection are largely protective, they do not guarantee effective immunity against subsequent infection.

This study leveraged a two-week USMC-mandated quarantine period during which baseline SARS-CoV-2 antibody status was established on arrival. Only baseline seronegative or seropositive participants who had multiple negative SARS-CoV-2 PCR tests during the supervised quarantine were included in the prospective study. The three negative PCR tests during quarantine helped ensure that infections diagnosed during basic training were not persistent infections but incident infection occurring during the prospective period. The two-week home quarantine preceding the supervised quarantine, as well as the relatively low frequency of infections diagnosed on arrival and during quarantine, further support that only incident infections were included in our analyses. The aggregate infection rate in both groups during the six weeks of observation at MCRDPI was 1,098 out of 2,436 (45.1%) participants. In contrast, only 28 of 3,249 (0.9%) participants were PCR positive on arrival at the supervised quarantine and less than 2% of participants became PCR positive during the two-week quarantine period. In view of the consecutive two periods of quarantine, the relatively low rate of infection during quarantine, and the three consecutive negative PCR tests, it is unlikely that any participants with persistent infection preceding their arrival at MCRDPI were entered into the prospective study. Even if this occurred, such cases would be unlikely to affect the relative risk calculations comparing the seropositive and seronegative groups. This methodology allowed for the creation of two well defined groups who entered basic training without active infection and differed primarily by baseline serology.

The two groups had similar demographic profiles, with the exception of a slightly higher prevalence in self-identified Hispanic and non-Hispanic black participants in the seropositive group. The seropositive group was almost 50% Hispanic and 22% non-Hispanic black participants compared to 22% and 12%, respectively in the seronegative group. This is likely due to minority populations having higher seroprevalence rates during the COVID-19 pandemic in general and among young adults specifically^27^.

Rates of infection and the risk reduction provided by seropositivity are important for understanding transmission dynamics for COVID-19, for epidemiologic modeling, and for estimating and achieving herd immunity levels, a major goal of mass vaccination strategies. Herd immunity is difficult to predict if the infection risk after natural and vaccine-induced immunity is unknown. Since SARS-CoV-2 vaccines may not provide sterile immunity, it is possible that both previously infected and vaccinated individuals may later become infected. It is not known if either can contribute to transmission events. We found only a modest, approximately 10-fold decrease in nares virus load as estimated by swab PCR Ct levels in the seropositive compared with the seronegative infected participants. This finding suggests that some reinfected individuals could have a similar capacity to transmit infection as those who are infected for the first time. The rate at which reinfection occurs after vaccines and natural immunity are important for estimating the percentage of the population that needs to be vaccinated to suppress the pandemic.

The crowded living conditions, demanding regimen and requirement for personal contact during basic training despite the pandemic leads not only to an increased risk for respiratory epidemics^29^, but also potentially to higher exposure levels. The close quarters and constant contact among recruits that are needed for team building allows a viral infection to rapidly proliferate within a unit. The physically and mentally demanding training environment may also suppress immunity. These conditions may contribute to the high infection rate we observed during the six-week study period. These factors are not typically present in the civilian community. Therefore, the study setting limits the generalizability of our findings to other settings where the frequency and intensity of exposure and the susceptibility of the host might differ.

The clinical outcomes between seropositive and seronegative groups were similar, with the majority (84.2% and 67.8%, respectively) being asymptomatic. The two groups did not show a significant difference in the duration of PCR positivity. No participants in either group needed inpatient care. Although our findings are limited to healthy young adults, studying this population does have the advantage of reducing the confounding factors of age and co-morbid illnesses^30^. Infection in seropositive participants was associated with lower SARS-CoV-2 IgG titers and absent or lower levels of neutralizing antibody activity. Young adults have high rates of asymptomatic and pauci-symptomatic infection, which has been associated with lower levels of antibodies and potentially a less robust immune memory response. This could lead to higher overall rates of re-infection among this population compared to other populations. We did not examine the role of cell mediated immunity or host, environment and virus factors leading to reinfection.

Since the study population is a fairly accurate representation of the races and ethnicities in the US population among 18-20-year-olds, the results are most applicable to young male adults. The relative risk of infection may be different in seropositive females and in adults of other ages or health status. Other limitations in our study include not being able to investigate the exposure event during a seropositive participant’s initial infection prior to arrival at quarantine, the inability to confirm initial SARS-CoV-2 infection by PCR in the seropositive group, and potentially missing detectable infections that occurred between biweekly sampling.

Our investigation is likely to underestimate the risk of SARS-CoV-2 infection in previously infected individuals because the seronegative group includes an unknown number of previously infected participants who did not have significant IgG titers in their baseline serum sample. Despite this underestimation, we found that previously infected participants identified by seropositivity are susceptible to repeat infection, with nearly one-fifth the incidence rate of those without evidence of previous infection. This suggests that COVID-19 vaccination may be necessary for control of the pandemic in previously infected young adults.

## Data Availability

Anonymized data will be made available on request.

## Contributors

AGL, IR, and SCS provided overall leadership and guidance to the investigation. YG performed statistical analysis and data curation. SV, CKP, CG, DLW, RL, and IR contributed to data analysis. RL, JM, EN, ESA, SM, NM, contributed to sample collection and management. CG, DLW, WDG contributed to data collection. SV, NAK, DE, AS, PB, NM, CM, DMP, SM, DS, PS performed serological assays. HWC, and VS performed PCR assays. AGL, SV, DLW, NAK, AS, RL, CM, CKP, MT, and IR contributed to data interpretation. AGL, CG, FJ, RL, EN supervised sample collection. AGL, CG, RL supervised data collection. RPT, FK, AB, and IR supervised serology assays. MS supervised PCR assays. MG and MT provided project administration and coordination. YG, SV, IR, CKP and VDN prepared figures. AGL, YG, SV, IR and SCS wrote and revised the original draft. All authors reviewed and approved the final manuscript.

## Acknowledgements

We thank Quanterix (Billerica, MA) for providing beta-level serological assay kits used for ELISA validation studies; Mary Anne Amper, Nitish Seenarine, Mital Vasoya, Illya Aronskyy and Braian Qela for outstanding technical assistance; German Nudelman and Stas Rirak for database development; Celia Gelernter and Adam B. Sealfon for editing assistance; Mitchel Rabinowitz for project management; Terri Brantley and Andrea Gates for administrative support; Adam Armstrong for strategic guidance; the many US Navy corpsmen who assisted in the logistics and sample acquisition; and the devoted Marine recruits who volunteered for this study. This work received funding from the Defense Health Agency through the Naval Medical Research Center (9700130) and from the Defense Advanced Research Projects Agency (contract number N6600119C4022).

## Supplementary Tables and Figures

**Supplementary Table 1:**
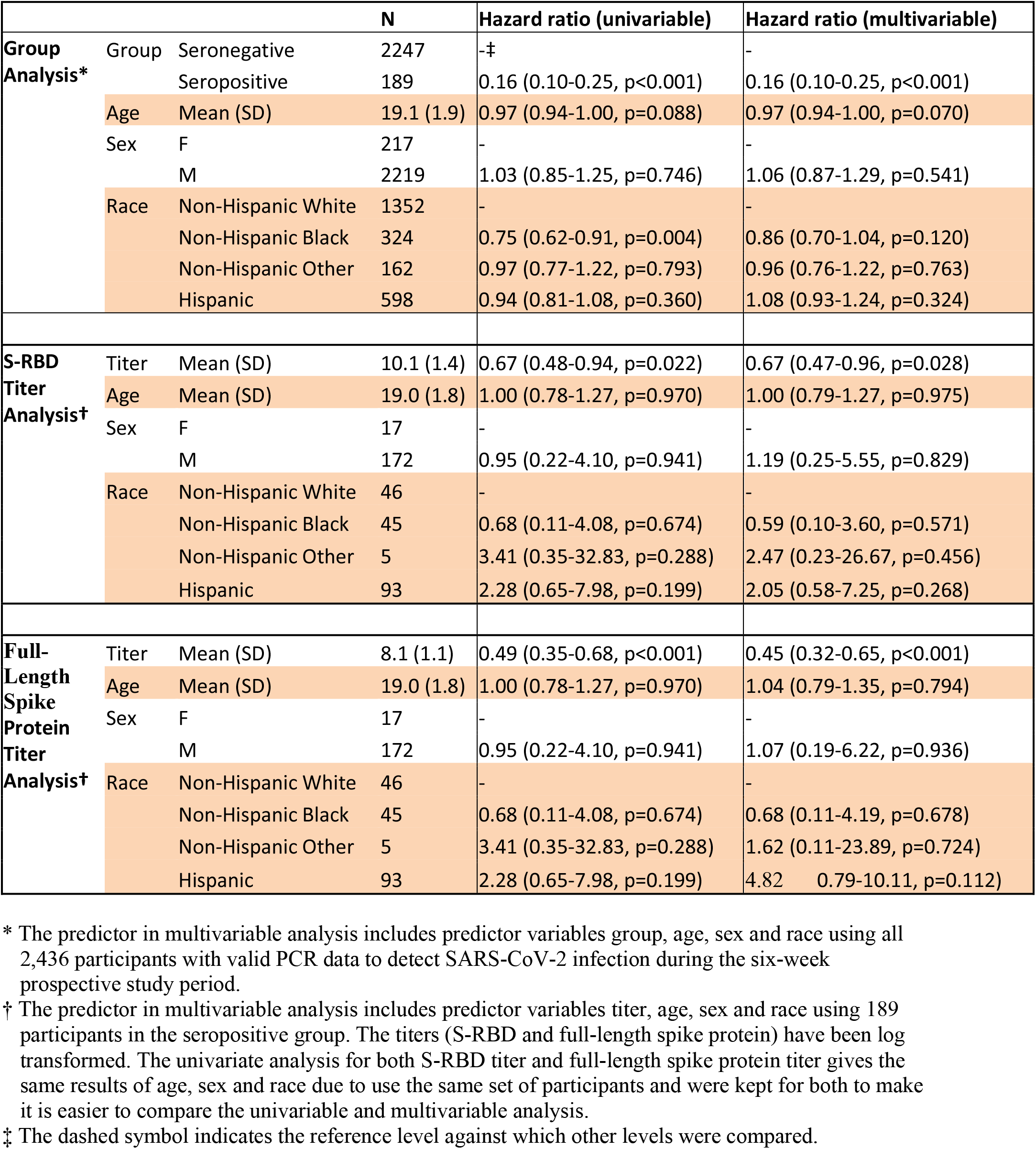
Cox proportional hazard regression analysis of group and titer association with SARS-CoV-2 infection.

**Supplementary Figure 1:**
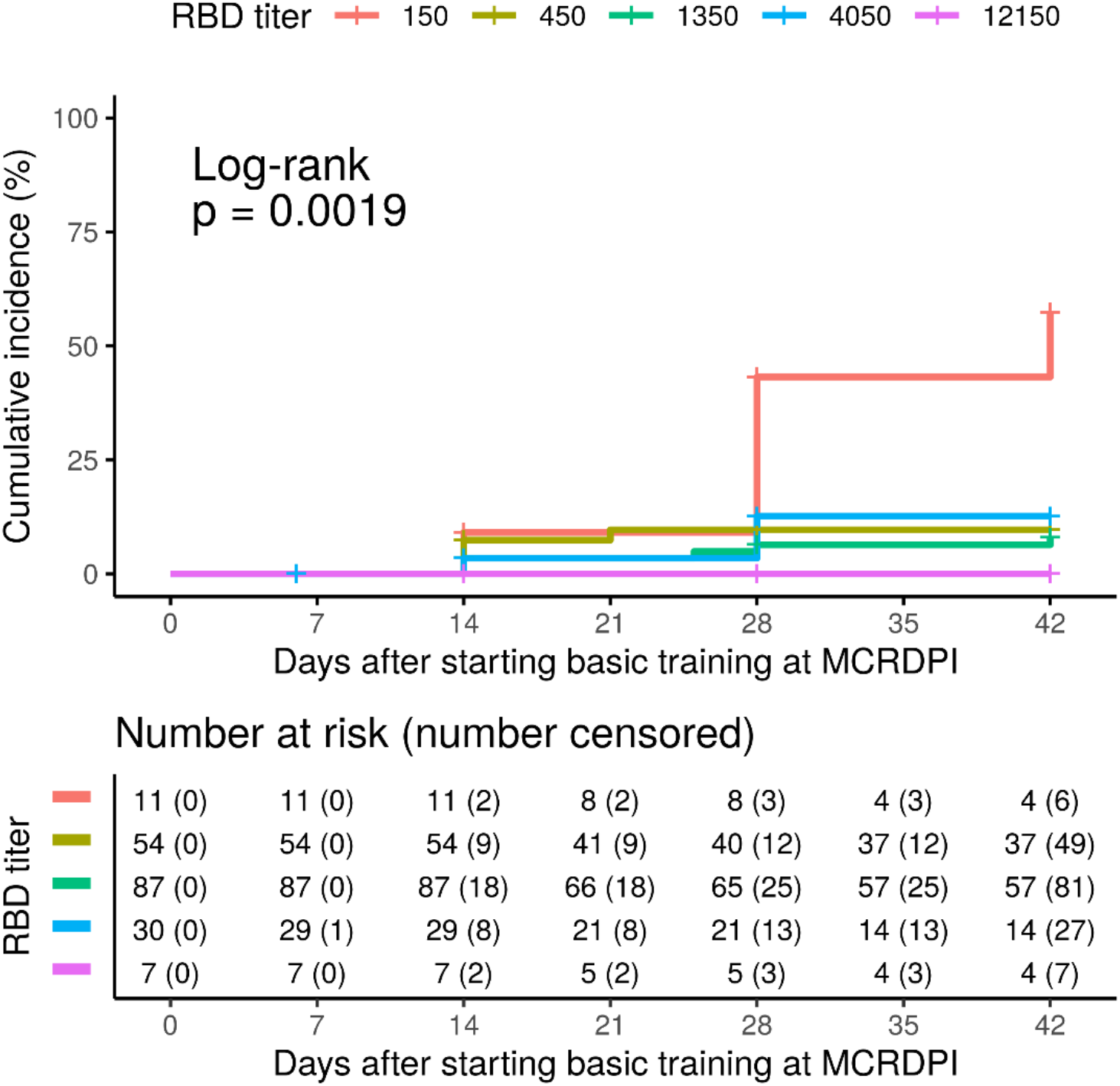
SARS-CoV-2 PCR positive incidence curves at different baseline RBD IgG titers during the six-week follow-up period. Cumulative incidence for testing PCR positive in the seropositive group at different baseline RBD IgG titers, which ranged from 1:150 to 1:12150. The numbers at risk and cumulated censored participants are listed below the table.

**Supplementary Figure 2:**
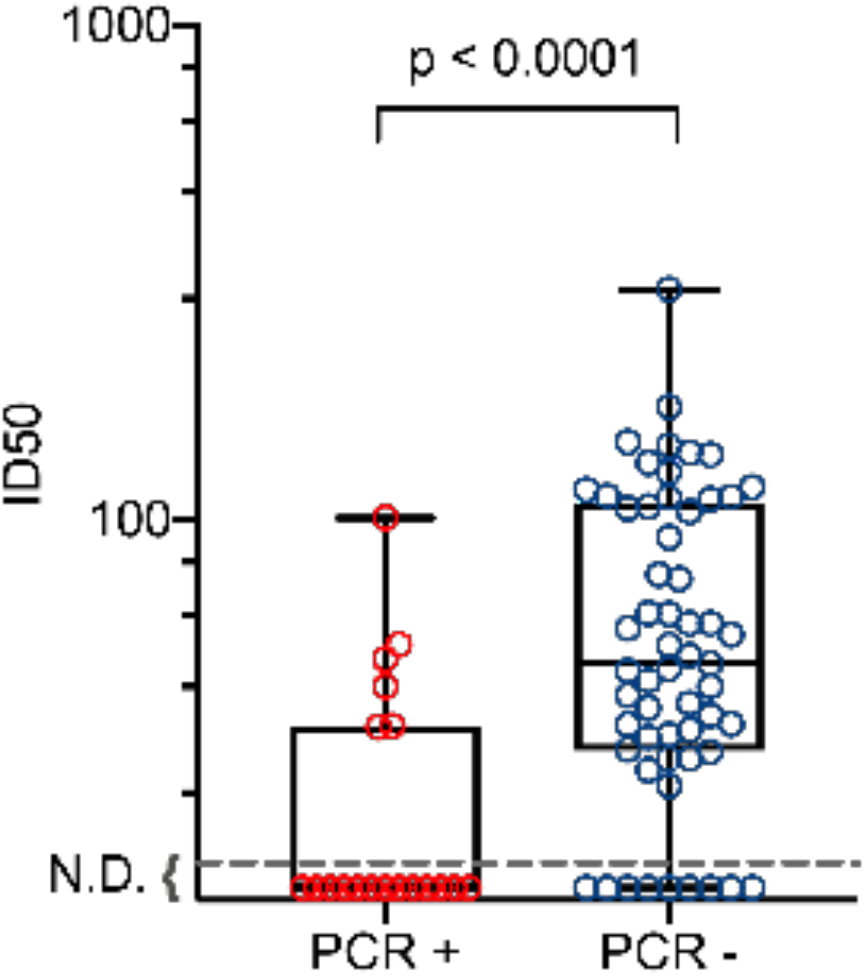
Baseline neutralizing antibodies in PCR+ and PCR-seropositive participants. The baseline 50% inhibitory dose (ID50) was determined for all seropositive participants who became SARS-CoV-2 PCR positive during the subsequent six-week observation period and in the first 54 seropositive participants enrolled who did not test PCR positive. N.D. indicates no inhibition detected at a 1:20 dilution which was the lowest titer evaluated. P value was assessed by the Cochran-Armitage test.

